# Early life blood pressure and cognitive function in mid/late life: A synthetic longitudinal cohort analysis

**DOI:** 10.64898/2026.02.24.26346790

**Authors:** Antonio J. Bustillo, Adina Zeki Al Hazzouri, M. Maria Glymour, Katrina L. Kezios

## Abstract

**BACKGROUND:** Hypertension is a well-studied dementia risk factor. The effects of hypertension on cognitive aging may vary by life stage, yet most prior studies have focused on mid- or late-life blood pressure. This gap reflects the lack of longitudinal cohorts spanning both childhood blood pressure measurements and later-life cognitive assessments. To address this gap, we examined the association between early-life blood pressure and mid/late-life cognition using a synthetic cohort approach.

**METHODS:** Using data fusion as a guiding framework, we pooled the Bogalusa Heart Study (BHS), which provides early-life blood pressure data (ages 4-16), with the CARDIA study, which provides mid/late-life cognitive data (ages 58-70), to create a synthetic longitudinal cohort. The cohorts overlapped between ages 17-57. Cognition comprised raw and z-scores measures of executive function, memory, and processing speed. Early life systolic blood pressure (SBP) was defined as a time-weighted average of SBP measurements between ages 4 and 16. We pooled data by exact- and distance-matching 10 BHS participants to each CARDIA participant on hypothesized mediators & confounders of the early life SBP and cognition relationship that overlapped in age of measurement between the two cohorts, including sociodemographics and ∼midlife measures of blood pressure, cognition, and vascular risk factors. Confounder-adjusted random intercept models were used to estimate the association between early-life SBP and cognitive outcomes.

**RESULTS:** There was no significant association between early life SBP and mid/late-life cognitive raw scores: executive function [β (95% CI) = 0.285 (−0.867, 1.437)], memory [−0.019 (−0.308, 0.269)], or processing speed [−0.188 (−1.611, 1.236]. Likewise, no associations were observed when using Z-scored cognitive outcomes.

**CONCLUSIONS:** We observed no evidence that early life SBP was associated with mid/late-life cognition, although interpretation of these findings depends on strong assumptions about cohort comparability and linkage validity that underpin the synthetic cohort approach. While our results lacked precision, we demonstrate a possible signal in the expected direction (i.e., greater systolic blood pressure results in worse cognitive functioning across all domains) that could be the focus of future work.

## INTRODUCTION

Hypertension affects about 1 in 2 adults (∼50%) aged 18 and over in the United States ^1^ and is a modifiable risk factor for cardiovascular disease and dementia ^2^ making it an important target for both prevention and intervention. However, as of 2023 the prevalence of hypertension in the United States remained well above the Healthy People 2030 target of 41.9% or lower,^1^ underscoring the importance of understanding its lifecourse health consequences and identifying critical periods for intervention.

Furthermore, hypertension and its associated morbidity and mortality are increasing in the United States among children and young adults ^3–5^, making it more critical than ever to understand the long-term consequences of elevated blood pressure beginning early in life, especially childhood. However, studying the later-life health effects of childhood blood pressure using a single cohort remains challenging because few cardiovascular cohorts have sufficient follow-up to measure both childhood blood pressure and outcomes occurring many decades later.

Hypertension may lead to changes in cognition via vascular mechanisms, such as small vessel disease ^6–9^ and may affect multiple cognitive domains, including executive function, memory, and attention.^10–14^ Although vascular mechanisms linking hypertension to cognitive impairment are well established, much less is known about whether the cognitive consequences of elevated blood pressure vary across the life course, particularly exposure in childhood. If hypertension-related vascular damage accumulates across the life course, elevated blood pressure beginning in childhood may contribute to pathological processes that influence cognitive aging long before clinical symptoms emerge. Despite this, the bulk of the epidemiological literature to date has largely focused on mid-life or late-life blood pressure or hypertension, showing that elevated mid-life blood pressure and hypertension diagnoses are associated with worse global cognition ^15–17^, memory ^15,16,18^, executive functioning ^15,16^, processing speed^19^, language ^19^, and risk of incident dementia^20,21^ in late life, with similar, though somewhat mixed findings for elevated late-life blood pressure and hypertension ^22–25^.

To our knowledge, no US-based cardiovascular cohort currently follows participants from childhood into later life while repeatedly measuring both blood pressure and cognitive function. Maintaining such a cohort—or newly establishing a birth cohort to eventually answer this question— presents logistical challenges and may be prohibitively time-consuming and resource-intensive. Alternatively, data fusion methods can be used to combine existing complementary data sources that separately provide early-life blood pressure exposure and mid/late-life cognitive outcomes. These complementary cohorts can be linked to create a synthetic longitudinal cohort that can be used to examine associations between early-life blood pressure and mid/late-life cognition during time frames otherwise impossible to examine in a single existing cohort.

Therefore, the aim of our study was to examine the association between early life blood pressure and mid/late-life cognition using a synthetic longitudinal cohort approach.

## METHODS

### Data sources

To create the synthetic cohort, we combined data from The Bogalusa Heart Study (BHS) and the Coronary Artery Risk Development in Young Adults Study (CARDIA) study. BHS provided early life blood pressure exposure data while CARDIA provided mid/late life cognition outcome data.

BHS is a community-based longitudinal study that examines the natural history of cardiovascular disease and its risk factors throughout the life course among residents of Bogalusa, Louisiana, with a series of cross-sectional surveys from childhood with ongoing data collection that commenced in 1973.^26–29^ This study used data collected between 1973-2016, including N=1298 individuals with cognitive data collected between ages 34 to 57. BHS includes prospectively collected measures of systolic & diastolic blood pressure beginning as early as age 4 and extending through age 57, making it well-suited for studying the long-term effects of early life blood pressure. Further, it has a rich array of cognitive measures collected between ages 34 and 57, spanning cognitive domains including global cognition, executive function, memory, language, and attention^27^ Because many of these cognitive domains overlap with those measured at similar ages in CARDIA, they provide important shared variables for linking participants across cohorts. Lastly, BHS includes sociodemographic variables, cardiovascular risk factors, other health-related variables, and healthcare utilization and insurance that can be leveraged for both cohort linkage and adjustment for potential confounding in the synthetic cohort **(Supplemental Table 1)**.

CARDIA is an ongoing multi-center prospective cohort study that investigates the development and risk factors for cardiovascular disease and has collected data from N=5115 community-dwelling adults across 4 metropolitan centers in the United States, including Birmingham, AL; Chicago, IL; Minneapolis, MN; and Oakland, CA.^30,31^ Baseline measurements were taken between 1985 and 1986 when participants were aged 17-30, and ongoing data collection has continued to the present day. We used 10 waves of data collected between 1985 and 2022 (Waves 0-10). CARDIA prospectively collects sociodemographic data, blood pressure, cardiovascular risk factors, and other health-related data at each wave between 1985 and 2022 (Waves 0-10) for individuals aged 17-70. Further, cognitive data was collected between 2010 and 2022 (Waves 8-10) among individuals aged 41 to 70 **(Supplemental Table 1)**.

### Blood pressure

Blood pressure data were repeatedly measured in both BHS and CARDIA. Blood pressures in BHS were measured by registered nurses using a mercury sphygmomanometer and averaged across 6 repeated measures.^32^ Blood pressures in CARDIA were measured 3 times by trained study personnel at 1-minute intervals after a 5-minute rest period using a Hawksley random zero sphygmomanometer between 1985-2006 (waves 0-8); after 2010 (waves 9 & 10), an Omron digital blood pressure monitor was used.^31,33^ Digital measures were calibrated to sphygmomanometer readings to avoid measurement bias.^31^ Our exposure of interest was mean early life systolic blood pressure (SBP), measured between ages 4-16 in the BHS and calculated as a time-weighted average.^34^

For matching, we created a variable for mean arterial pressure (MAP) from blood pressure measures collected during the matching time period (approximately midlife). MAP was calculated as (1/3*SBP + 2/3*DBP) and dichotomized into low and high groups using a median split.

### Cognition

We obtained cognitive outcome data for individuals aged 58-70 from CARDIA, measured at three study visits from 2010-2022 (waves 8, 9, & 10). Mid/late-life cognition (ages 58-70) was defined using cognitive domain scores, including executive function, memory, and processing speed, from all waves of CARDIA data measured between ages 58-70.

Executive function (ages 58-70) was defined using the Stroop interference task score from CARDIA.^35–37^ The Stroop interference task measures the time required to respond to one stimulus while suppressing another; higher scores indicate worse executive functioning.^38^

Memory (ages 58-70) was defined using the Rey Auditory Verbal Learning Test (RAVLT)^35,37^ available in CARDIA. RAVLT measures verbal memory; scores reflect the number of words recalled, with higher scores indicating better memory.^38^ Memory in BHS was defined analogously using the delayed free recall subtest from the Wechsler Memory Scale, 4^th^ edition.^27^

Processing speed (ages 58-70) was analogously defined in CARDIA using the Digit Symbol Substitution Test (DSST) ^35,36,39^. The task involves coding symbols to numbers under a 2-minute time constraint.

Higher scores, which reflect more correctly coded symbols, indicate better processing speed ^38^Processing speed was analogously defined in BHS was the Digit Symbol Coding Test (DSC), which involved the same task.^27^ Cognitive domain scores were calculated as both raw and z-transformed scores, with z-transformed scores normed to individuals aged 58-70.

The earliest cognitive assessment available during the period of age overlap between the cohorts (approximately midlife) was used for matching, while later cognitive assessments (i.e., at ages 58-70 that are uniquely represented in CARDIA) served as the analytic outcomes of interest. Like intermediate measures of blood pressure, earlier cognitive performance was included as a matching variable because it represents an intermediate life course measure shared by both cohorts that helps preserve associations between the exposure and later cognitive outcomes after linkage.

### Statistical analyses

Our target sample consisted of CARDIA participants with cognitive assessments available beginning in 2010 (mean age 50 years). These participants were matched to BHS participants who completed cognitive testing around the same period (2013-2016) and were of similar age (mean age 48 years). This sampling strategy minimized differences in participant age and calendar time between BHS and CARDIA during the matching period.

Statistical analyses proceeded in four broad steps: (1) selection of matching variables, (2) data preparation, (3) data pooling, and (4) effect estimation.

Matching variables were selected based on their hypothesized role in preserving the relationship between early life blood pressure and later-life cognition that would have been observed had both variables been measured in the same individuals. Such variables included potential mediators or confounders along hypothesized pathways linking early life blood pressure and later cognition,^40,41^ measured at approximately the same participant age and calendar year in both cohorts. These variables included age, sex (male, female), race (Black, White), executive function, memory, processing speed, SBP, diastolic blood pressure (DBP), low/high MAP, low-density lipoprotein (LDL), education (less than high school, high school, some college, Bachelor’s degree or greater), and ever smoked (yes, no). Exact matching on low/high MAP improved balance between cohorts with respect to SBP and DBP, two key linking variables. In CARDIA, these matching variables were measured in 2010 between ages 42-59, and in BHS they were measured between 2013-2016 among participants aged 35-57 **(Supplemental Figure 1)**.

Data preparation involved harmonizing and imputing missing data for the selected matching variables. Harmonization for most variables involved re-coding variables from either cohort to conform to a common coding scheme. Harmonization for cognition was achieved by creating cognitive domain Z-scores normed within each cohort. Imputation for missing data was carried out using the last-value-carried-forward/back method for categorical variables (including education & smoking); no data were imputed for those completely missing smoking and education history. Missing data for continuous matching variables were imputed using multilevel joint imputation with random intercepts and random slopes for exam year, implemented in the *panImpute* R package.^42^ The data preparation step concluded with the construction of a complete-case dataset to exclude cases completely lacking smoking & education history. For CARDIA, we excluded this N=176 (∼5%) and for BHS we excluded N=26 (∼2%). This complete case dataset comprised the matching pool for both cohorts **(Figure 1).**

**Figure 1.**
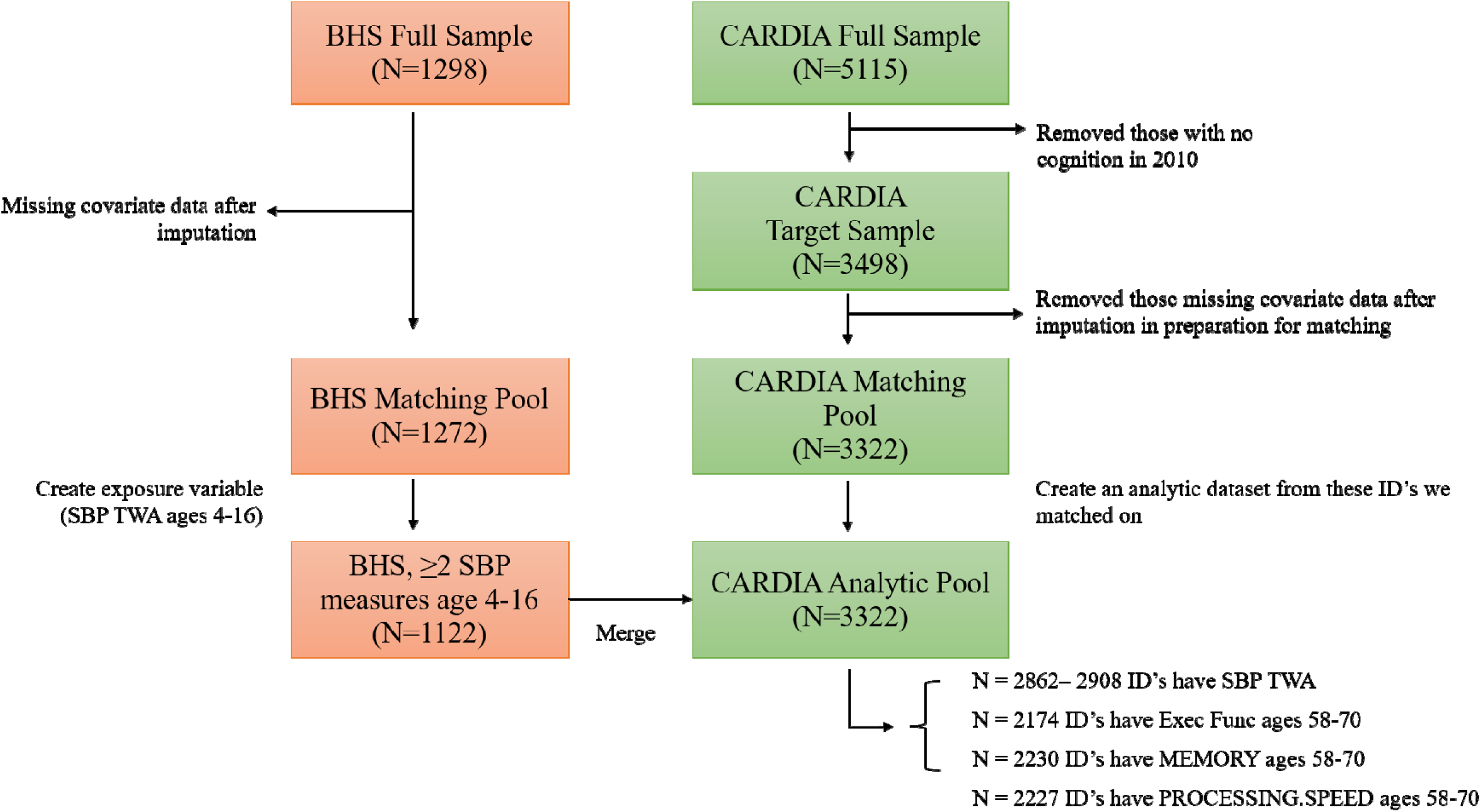
Sample size flow chart.

We used a previously described matching-based method to link participants across cohorts on the aforementioned harmonized matching variables.^41^ The goal was to match each individual with mid/late-life cognition at ages 58-70 in CARDIA to comparable individuals with early-life blood pressure exposure data from BHS (ages 4-16). Matching was conducted using the *matchIt* R package.^43^ We exact-matched on race, sex, education, and low/high MAP. Distance-matching was conducted on memory, executive function, processing speed, SBP, DBP, BMI, LDL and smoking using the Malahanobis distance. We matched many-to-one (10 BHS:1 CARDIA) with replacement.^43^ To ensure every CARDIA participant received 10 matches, we allowed each BHS participant to be reused up to 89 times, the minimum reuse limit required to achieve 10 matches for all CARDIA participants.^44^ Covariate balance was assessed graphically using Love plots.

Because this matching-based approach is procedurally similar to hot deck imputation, ^40,45^ uncertainty in the early life blood pressure values assigned to each CARDIA participant through matching should be accounted for. Matched cases were thus randomly sampled into 10 datasets. Within each dataset, we ran random intercept models to estimate the effect of mean early life SBP on mid/late life cognition. Models were adjusted for age, sex, race, education, and smoking. Separate models were run for each cognitive domain. Results for each model were averaged across the 10 datasets, and variances were calculated using Rubin’s rules as the sum of within and between matched datasets.^46^ Estimates were pooled using *mitools* R package.^47^

## RESULTS

We used a sample size of N=3498 CARDIA participants and N=1298 BHS participants. **Table 1** shows sample descriptives by component study cohort prior to matching for variables in CARDIA measured in 2010 between ages 42-59 and BHS measured between 2013-2016 between ages 35-57. In both cohorts, participants were predominantly middle-aged (median (Q1, Q3) age was 51 (47, 53) for CARDIA and 49 (44, 53) for BHS) was majority female, and White. Participants in CARDIA were more likely to have possessed a bachelor’s degree or higher, while BHS participants were more likely to have ever smoked.

**Table 1.** Descriptive statistics by cohort.

| **Characteristic** | **BHS** | **CARDIA** |
| --- | --- | --- |
|  | N = 1,298 | N = 3,498 |
| Age (years)<sup>a</sup> | 49 (44, 53) | 51 (47, 53) |
| Sex<sup>b</sup> |  |  |
| Female | 764 (59%) | 1,981 (57%) |
| Male | 534 (41%) | 1,517 (43%) |
| Race<sup>b</sup> |  |  |
| Black | 447 (35%) | 1,640 (47%) |
| White | 848 (65%) | 1,858 (53%) |
| Education<sup>b</sup> |  |  |
| Less than High School | 32 (2.5%) | 317 (9.1%) |
| High School | 630 (49%) | 1,122 (32%) |
| Some College | 524 (40%) | 437 (12%) |
| Bachelor's Degree or higher | 112 (8.6%) | 1,622 (46%) |
| Employed<sup>b</sup> | 813 (63%) | 2,750 (79%) |
| Birth Year<sup>a</sup> | 1966 (1962, 1970) | 1960 (1957, 1963) |
| Exam Year<sup>b</sup> |  |  |
| 2010 | 0 (0%) | 3,498 (100%) |
| 2013 | 68 (5.2%) | 0 (0%) |
| 2014 | 563 (43%) | 0 (0%) |
| 2015 | 605 (47%) | 0 (0%) |
| 2016 | 62 (4.8%) | 0 (0%) |
| Systolic blood pressure (mmHg)<sup>a</sup> | 121 (112, 133) | 118 (109, 128) |
| Diastolic blood pressure (mmHg)<sup>a</sup> | 78 (71, 85) | 74 (67, 82) |
| Body mass index (kg/m<sup>2</sup>)<sup>a</sup> | 30 (26, 36) | 29 (25, 34) |
| Low density lipoprotein (mg/dL)<sup>a</sup> | 112 (90, 135) | 110 (89, 132) |
| Ever smoked<sup>b</sup> |  |  |
| Yes | 826 (64%) | 1,931 (55%) |
| No | 391 (30%) | 1,567 (45%) |
| Missing | 81 (6.2%) | 0 (0%) |
| Processing Speed Z-score<sup>a</sup> | 0.01 (-0.72, 0.69) | 0.00 (-0.68, 0.68) |
| Memory Z-score<sup>a</sup> | 0.00 (-0.68, 0.68) | -0.10 (-0.71, 0.82) |
| Executive Function Z-score<sup>a</sup> | -0.24 (-0.63, 0.40) | -0.16 (-0.62, 0.39) |

<sup>a</sup>Median (Q1, Q3)<sup>b</sup>N (%)

In the pre-matched pool, each cohort was balanced on several characteristics used for matching including memory, executive function, & processing speed Z-scores, LDL, and sex. **Table 2** details the standardized mean difference (SMD) for all matching covariates before and after matching. SBP, DBP, BMI, smoking, education, and race showed greater imbalance (i.e., absolute SMD > 0.1) prior to matching. Matching created exact or improved balance across all these covariates **(Table 2 & Figure 2).** Large SMD (i.e., > 0.1) persisted before and after matching (i.e., SMD > 0.1); however, this difference translated to 0.8-2 years.

**Figure 2.**
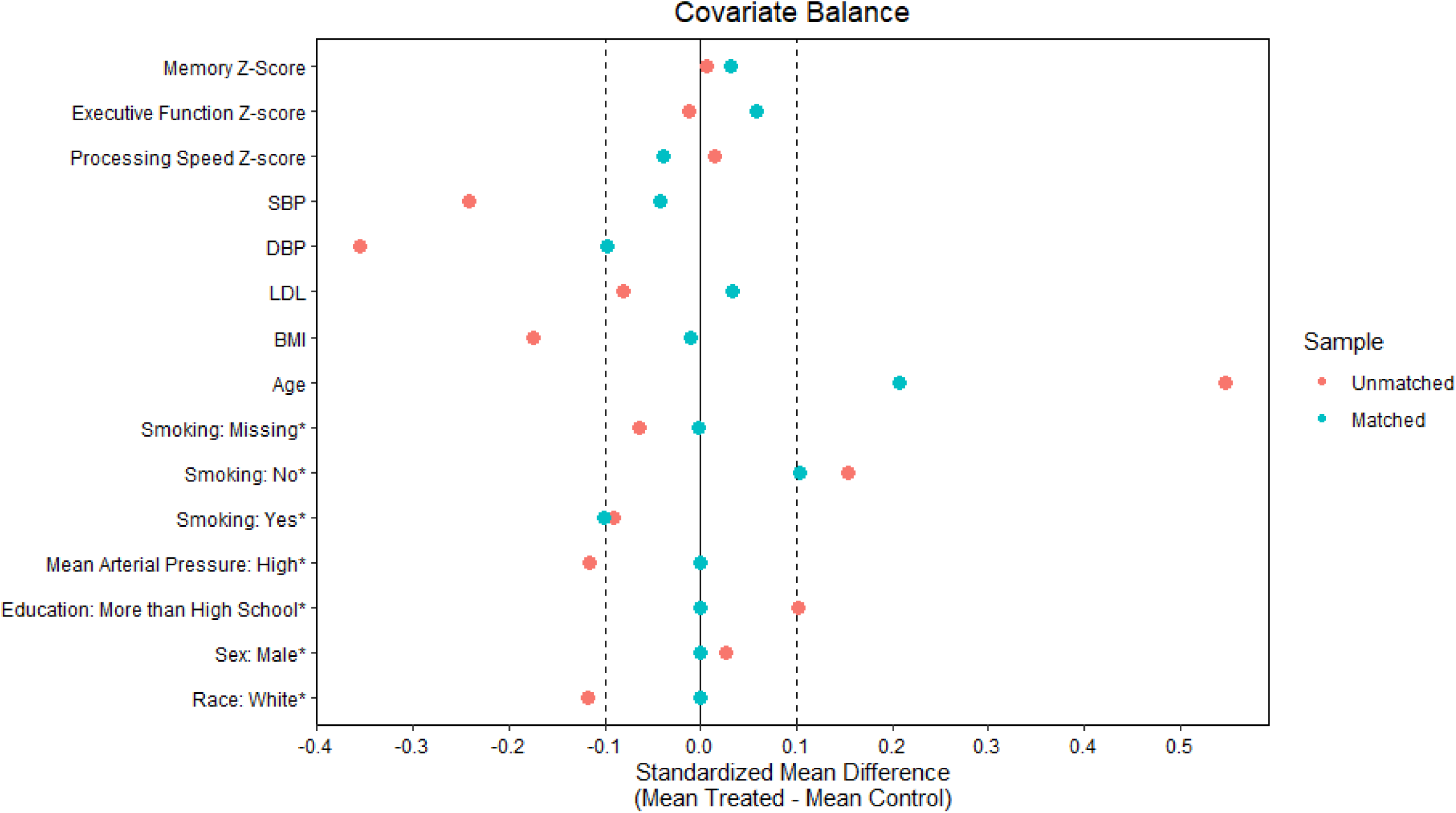
Love plot depicting covariate balance post-matching.

**Table 2.** Means and standardized mean difference for each covariate by cohort pre- & post-matching.

| **Covariate** | **Pre-Matching** |  |  | **Post-Matching** |  |  |
| --- | --- | --- | --- | --- | --- | --- |
|  | **CARDIA**<br>**(N=3322)** | **BHS**<br>**(N=1272)** | **SMD<sup>a</sup>** | **CARDIA**<br>**(N=3322)** | **BHS**<br>**(N=1272)** | **SMD<sup>a</sup>** |
| Memory Z-Score | 0.01 | 0.003 | 0.007 | 0.01 | -0.020 | 0.031 |
| Executive Function Z-Score | -0.002 | 0.009 | -0.011 | -0.002 | -0.061 | 0.059 |
| Processing Speed Z-Score | 0.011 | -0.003 | 0.014 | 0.011 | 0.049 | -0.039 |
| SBP | 119.534 | 123.417 | -0.242 | 119.534 | 120.208 | -0.042 |
| DBP | 74.745 | 78.719 | -0.355 | 74.745 | 75.84 | -0.098 |
| LDL | 111.826 | 114.456 | -0.081 | 111.826 | 110.749 | 0.033 |
| BMI | 30.142 | 31.405 | -0.175 | 30.142 | 30.219 | -0.011 |
| Age | 50.156 | 48.167 | 0.547 | 50.156 | 49.399 | 0.208 |
| Smoking: Yes | 0.546 | 0.636 | -0.090 | 0.546 | 0.646 | -0.100 |
| Smoking: No | 0.454 | 0.3 | 0.153 | 0.454 | 0.351 | 0.103 |
| Smoking: Yes | 0.546 | 0.636 | -0.090 | 0.546 | 0.646 | -0.100 |
| Mean Arterial Pressure: High | 0.473 | 0.588 | -0.115 | 0.473 | 0.473 | 0 |
| Education: More than High School | 0.595 | 0.493 | 0.102 | 0.595 | 0.595 | 0 |
| Sex: Male | 0.435 | 0.409 | 0.026 | 0.435 | 0.435 | 0 |
| Race: White | 0.541 | 0.658 | -0.117 | 0.541 | 0.541 | 0 |

<sup>a</sup>SMD = Standardized mean difference; calculated as the difference between CARDIA and BHS.

**Table 3** shows the estimated associations and 95% confidence intervals for the mean early life SBP (ages 4-16) with each cognitive domain. Estimates were positive for executive function and negative for memory and processing speed, suggesting that each 10 mmHg increase in mean early SBP led to worse cognitive performance across each domain. However, unadjusted estimates became attenuated after adjusting for age, race, sex, education, & smoking measured at mid-life and all estimated lacked statistical precision. No significant association was seen for any raw cognitive domain score nor for Z-scores.

**Table 3.** Association between early life SBP and mid/late life cognitive function in a synthetic longitudinal cohort.

|  | **Model 1<sup>c</sup>** | **Model 2<sup>d</sup>** | **Model 3<sup>e</sup>** |
| --- | --- | --- | --- |
| **Cognitive Domain (Raw Scores)<sup>a</sup>** | **Estimate (95% CI)<sup>f</sup>** | **Estimate (95% CI)<sup>f</sup>** | **Estimate (95% CI)<sup>f</sup>** |
| Executive Function | 0.636 (-0.409, 1.682) | 0.272 (-0.879, 1.422) | 0.260 (-0.906, 1.425) |
| Memory | -0.139 (-0.455, 0.176) | -0.012 (-0.297, 0.273) | -0.012 (-0.30, 0.274) |
| Processing Speed | -0.816 (-2.433, 0.8) | -0.156 (-1.567, 1.255) | -0.136 (-1.560, 1.288) |
| **Cognitive Domain (Z-Scores)<sup>b</sup>** | **Estimate (95% CI)<sup>f</sup>** | **Estimate (95% CI)<sup>f</sup>** | **Estimate (95% CI)<sup>f</sup>** |
| Executive Function | 0.638 (-0.406, 1.681) | 0.023 (-0.074, 0.12) | 0.021 (-0.076, 0.120) |
| Memory | -0.04 (-0.129, 0.05) | -0.003 (-0.084, 0.077) | -0.003 (-0.003, 0.078) |
| Processing Speed | -0.05 (-0.148, 0.049) | -0.01 (-0.095, 0.076) | -0.008 (-0.10, 0.078) |

<sup>a</sup>Cognitive raw scores between ages 58-70. Executive function measured using the Stroop interference task; memory measured using the Rey Verbal Auditory Learning Test; processing speed measured using the Digit Symbol Substitution Test.
<sup>b</sup>Cognitive Z-scores between ages 58-70. Scores were normed to individuals in CARDIA with cognition between ages 58-70.
<sup>c</sup>Model 1 unadjusted.
<sup>d</sup>Model 2 adjusted for age, race, sex, & education.
<sup>e</sup>Model 3 adjusted for age, race, sex, education, & smoking.
<sup>f</sup>Estimate (95% CI) for each 10 mmHg increase in the time-weighted average in systolic blood pressure between ages 4-16.

## CONCLUSIONS

Our study observed that, after pooling two US-based cardiovascular cohorts, there was no association between early life SBP and mid/late-life cognition. While estimates trended in the expected direction for the association between systolic blood pressure and cognition at other life stages, estimates lacked precision. Our results add to the body of evidence that examines blood pressure exposure throughout the lifecourse and its association with cognition in later life stages. Specifically, our findings suggest that early life systolic blood pressure may have a smaller (or null) influence on later cognitive function compared with blood pressure measured at later ages, although additional studies are needed.

Although the associations were not statistically significant, all point estimates were consistent with the hypothesized adverse effect of higher blood pressure in earlier life on later cognitive outcomes. For example, prior studies in CARDIA by Yaffe et al (2014) showed that elevated blood pressure throughout early adulthood (ages 18-39) was associated with worse verbal memory, processing speed, and executive functioning after 25 years of follow-up.^48^ Taken together, these findings suggest that blood pressure measured during childhood and adolescence (i.e., ages 4-17 in our study) may have a smaller influence on mid/later-life cognitive function than blood pressure during early or mid-adulthood. However, the interpretation of these findings depends on strong assumptions of cohort comparability and linkage validity that underpin the synthetic cohort approach, and the findings should be replicated in other single-cohort or synthetic cohort settings.

A major strength of this study was the availability of numerous shared variables spanning hypothesized mediating and confounding pathways between early life blood pressure and later-life cognition.

Importantly, these included intermediate (∼midlife) measures of the exposure (blood pressure) and outcome (cognitive function), which previous work has shown to serve as particularly informative linking variables.^41,49^ Additionally, we observed substantial overlap in SBP across the life course, from early life to mid-life, enhancing our ability to achieve close matches on this key variable, and the cohorts shared a rich array of cardiovascular risk factors to capture important front- and back-door pathways linking the exposure and outcome. Further, while there was little overlap in the specific cognitive tests, the battery in each cohort was sufficiently rich to allow calculation of comparable cognitive domains. Finally, our pooling methodology is rooted in a causal data-fusion framework and supported by simulation studies, providing principled guidance for constructing the synthetic cohort and estimating the association of early life blood pressure and mid/late-life cognition within it.

This study has some notable limitations. Most importantly, the validity of our findings rests on key assumptions regarding the comparability of the cohorts and the sufficiency of the linking variables used to construct the synthetic cohort. We sought to maximize cohort comparability as best possible by linking two US-based cardiovascular cohorts from overlapping geographic regions, matching participants of similar ages, and selecting variables for matching that were measured during similar calendar periods.

Additionally, although we cannot rule out the possibility that important linking pathways were omitted, which could bias synthetic cohort estimates,^41^ both cohorts included a rich set of shared variables that we believe support valid linkage. Additionally, missing data in the matching variables reduced the pool of eligible participants, requiring imputation of covariates prior to linkage and potentially introducing additionally uncertainty in our estimates. Finally, because there was limited overlap in the specific cognitive tests between the two cohorts, the midlife cognition variables used for matching were harmonized using cognitive domain Z-scores rather than identical tests. Future work should further evaluate the comparability of the underlying latent cognitive constructs in each cohort to strengthen harmonization and therefore matching between BHS and CARDIA.

Our study has some notable public health implications. First, our study helps us understand whether early life blood pressure should form part of the early life exposome, which leads to poorer cognitive aging in mid/late life. While our results lacked precision, we demonstrate a possible signal in the expected direction (i.e., greater systolic blood pressure results in worse cognitive functioning across all domains) that could be the focus of future work.

Future studies that aim to examine the associations between early life exposure & outcomes in later life should aim to use data sources with greater overlap in all outcomes. Additionally, as the early cohort continues to age, future analysis should focus on confirming our findings in the BHS cohort. Finally, future analyses should examine the effects of other vascular exposures in early life, including diastolic blood pressure, pulse pressure, and heart rate.

## Data Availability

All data produced in the present study are available upon reasonable request to the authors.

