## Supplemental Table 1 for "Early life blood pressure and cognitive function in mid/late life: A synthetic longitudinal cohort analysis"

| **BHS (Early Life Cohort) Characteristics** | | | | | |
| --- | --- | --- | --- | --- | --- |
| Years of data collection | 1973-2016 | | | | |
| Follow-up frequency | Every 2-3 years on average | | | | |
| Number of waves/study visits | 3 to 16 study visits | | | | |
| Age range (across waves) | 4-57 years | | | | |
| Age of measurement | **4-16** | **17-34** | **35-44** | **45-57** | **58-70** |
| Blood Pressure | ✔ | ✔ | ✔ | ✔ |  |
| Sociodemographics | ✔ | ✔ | ✔ | ✔ |  |
| Cardiovascular risk factors | ✔ | ✔ | ✔ | ✔ |  |
| Cognition |  |  | ✔ | ✔ |  |
| **CARDIA (Mid/Late life Cohort) Characteristics** | | | | | |
| Years of data collection | 1985-2022 | | | | |
| Follow-up frequency | Every 2 to 5 years | | | | |
| Number of waves/study visits | 10 waves | | | | |
| Age range (across waves) | 17-70 years | | | | |
| Age of measurement | **4-16** | **17-34** | **35-44** | **45-57** | **58-70** |
| Blood Pressure |  | ✔ | ✔ | ✔ | ✔ |
| Sociodemographics |  | ✔ | ✔ | ✔ | ✔ |
| Cardiovascular risk factors |  | ✔ | ✔ | ✔ | ✔ |
| Cognition |  |  | ✔ | ✔ | ✔ |

**Supplemental Table 1.** Study characteristics by cohort
