## Supplementary figures and images for "Early life blood pressure and cognitive function in mid/late life: A synthetic longitudinal cohort analysis"

### Supplemental Figure 1

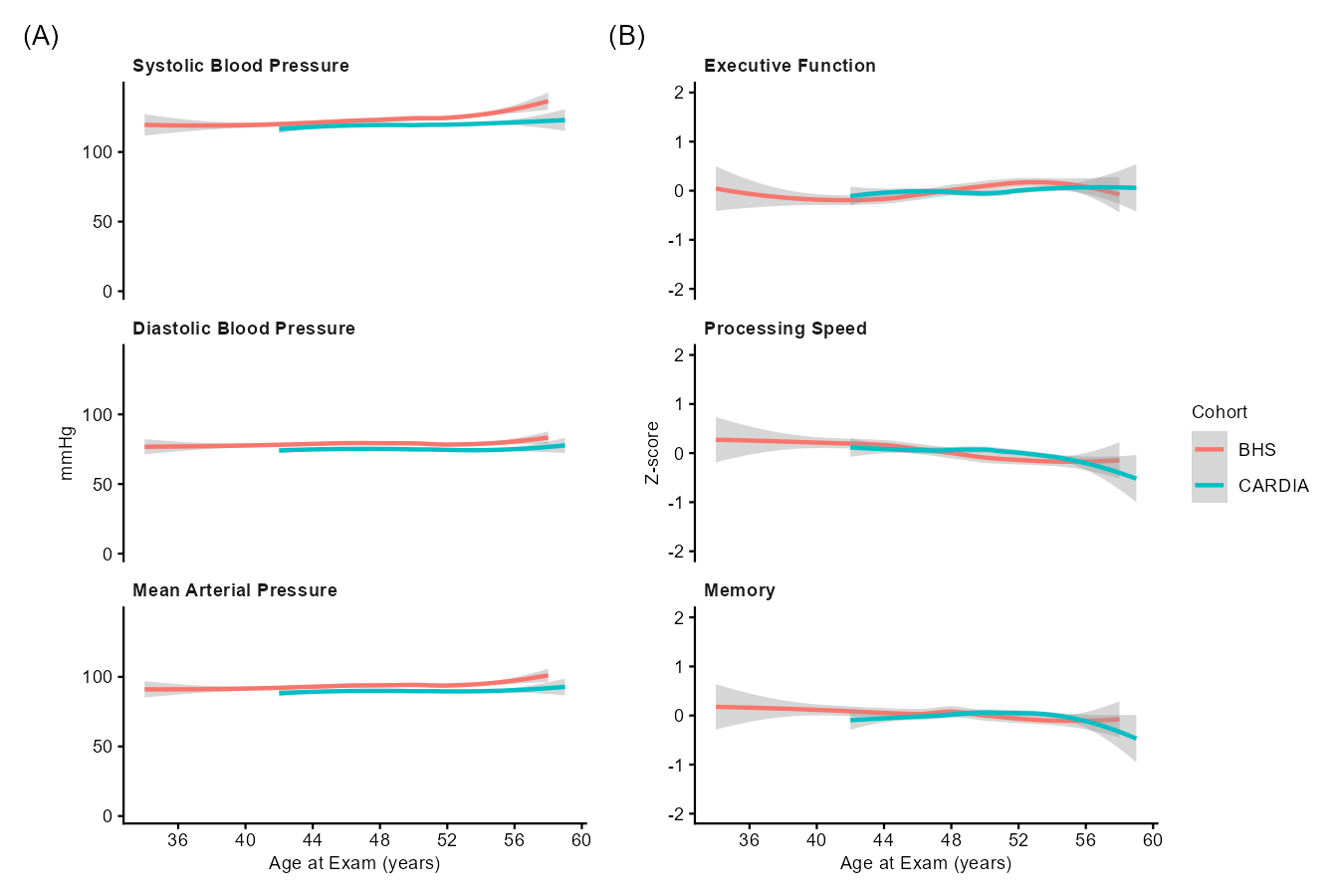

**Supplemental Figure 1.** Overlap in blood pressure variables & cognition pre-matching.
